# Use of whole genome sequencing to estimate the contribution of immune evasion and waning immunity to decreasing COVID-19 vaccine effectiveness during alpha and delta variant waves

**DOI:** 10.1101/2022.08.25.22278443

**Authors:** Margaret L Lind, Richard Copin, Shane McCarthy, Andreas Coppi, Fred Warner, David Ferguson, Chelsea Duckwall, Ryan Borg, M Catherine Muenker, John Overton, Sara Hamon, Anbo Zhou, Derek AT Cummings, Albert I. Ko, Jennifer D Hamilton, Wade Schulz, Matt T. Hitchings

**Affiliations:** Department of Epidemiology of Microbial Diseases, Yale School of Public Health, New Haven, CT, USA; Regeneron Pharmaceuticals, Inc, Tarrytown, NY, USA; Section of Cardiovascular Medicine, Department of Internal Medicine, Yale School of Medicine, New Haven, CT, USA; Center for Outcomes Research and Evaluation, Yale-New Haven Hospital, New Haven, CT, USA; Department of Biology, University of Florida, Gainesville, FL, USA; Emerging Pathogens Institute, University of Florida, Gainesville, FL, USA; Instituto Gonçalo Moniz, Fundação Oswaldo Cruz, Salvador, BA, Brazil; Department of Internal Medicine, Yale School of Medicine, New Haven, CT, USA; Department of Laboratory Medicine, Yale University School of Medicine, New Haven, CT, USA; Department of Biostatistics, College of Public Health and Health Professions & College of Medicine, University of Florida, Gainesville, FL, USA

**Keywords:** Vaccine effectiveness, COVID-19, whole genome sequencing, waning, variant specific immune evasion

## Abstract

**Background:** The decline in COVID-19 mRNA vaccine effectiveness (VE) is well established, however the impact of variant-specific immune evasion and waning protection remains unclear. Here, we use whole-genome-sequencing (WGS) to tease apart the contribution of these factors on the decline observed following the introduction of the Delta variant. Further, we evaluate the utility of calendar-period-based variant classification as an alternative to WGS.

**Methods:** We conducted a test-negative-case-control study among people who received SARS-CoV-2 RT-PCR testing in the Yale New Haven Health System between April 1 and August 24, 2021. Variant classification was performed using WGS and secondarily by calendar-period. We estimated VE as one minus the ratio comparing the odds of infection among vaccinated and unvaccinated people.

**Results:** Overall, 2,029 cases (RT-PCR positive, sequenced samples) and 343,985 controls (negative RT-PCRs) were included. VE 14-89 days after 2^nd^ dose was significantly higher against WGS-classified Alpha infection (84.4%, 95% confidence interval: 75.6-90.0%) than Delta infection (68.9%, CI: 58.0-77.1%, p-value: 0.013). The odds of WGS-classified Delta infection were significantly higher 90-149 than 14-89 days after 2^nd^ dose (Odds ratio: 1.6, CI: 1.2-2.3). While estimates of VE against calendar-period-classified infections approximated estimates against WGS-classified infections, calendar-period-based classification was subject to outcome misclassification (35% during Alpha period, 4% during Delta period).

**Conclusions:** These findings suggest that both waning protection and variant-specific immune evasion contributed to the lower effectiveness. While estimates of VE against calendar-period-classified infections mirrored that against WGS-classified infections, our analysis highlights the need for WGS when variants are co-circulating and misclassification is likely.

**Summary of main points:** Using whole genome sequencing, we provide direct evidence of waning vaccine effectiveness and variant-specific immune evasion during the Delta wave. Effectiveness estimates against calendar-period-classified infections approximated estimates against WGS-classified infections, however, calendar-period classification was associated with a variant misclassification.

## Introduction

As the number of cases and deaths among vaccinated and unvaccinated people rose following the introduction of the Delta variant of SARS-CoV-2 in the summer of 2021, concerns regarding variant specific immune evasion surfaced.[1,2] While some studies found vaccine effectiveness against Delta was lower than previously circulating variants (such as Alpha)[3–11], other studies found the differences were likely driven by waning levels of protection, not variant-specific immune evasion.[12–17]

However, interpreting and comparing studies of vaccine effectiveness is challenging due to the use of inconsistent variant-specific outcome definitions. In the absence of whole genome sequencing (WGS), studies commonly rely on calendar-period-based variant classification.[6,8,10,11,15,18,19] Though studies reliant on this approach have incorporated sensitivity analyses with varying time periods, this approach remains subject to unaccountable outcome (variant) misclassification. Despite its frequent use, a comparison of vaccine effectiveness resulting from WGS and calendar-period-based variant classification has not been completed. As a result, the reliability of calendar-period-based estimates remains uncertain.

In this retrospective analysis, we leveraged data from a large cohort of individuals enrolled in the Yale New Haven Health System (YNHH) who underwent RT-PCR testing to estimate and compare the effectiveness of mRNA vaccines (mRNA-1273-Moderna and BNT162b2-Pfizer) against infection, symptomatic infection and COVID-19 associated hospitalization with WGS-classified Alpha, Delta, and other co-circulating variants. Further, we examined whether protection conferred by primary series vaccination declined over time since series completion (second dose receipt). Finally, to examine the reliability of calendar-period-based variant classification, we compared estimates of vaccine effectiveness during the calendar period of Alpha and Delta variant predominance with those obtained from WGS.

## Methods

### Study Design and Setting

We conducted a test-negative-case-control (TNCC) analysis using RT-PCR tests collected between April 1 and August 24, 2021, as part of the larger Studying COVID-19 Outcomes after SARS-CoV-2 Infection and Vaccination (SUCCESS) Study at YNHH. The YNHH is a large academic health system comprising four delivery networks in Connecticut, southeastern New York, and western Rhode Island. We chose a TNCC design because it mitigates the risk of confounding introduced by care-seeking and testing access and has been shown to provide estimates of vaccine effectiveness consistent with those from randomized control trials.[20–24]

### RT-PCR Collection and Sequencing

SARS-CoV-2 RT-PCR testing was broadly implemented at YNHH to screen people with suspected SARS-CoV-2 exposure and patients who underwent procedures or were admitted for hospitalization. Specimens were collected using nasopharyngeal/oropharyngeal swabs and primarily evaluated with QuantStudio 7 Flex Real-Time PCR System (Applied Biosystems, Waltham, MA), BHG Probe Panther (Hologic, Marlborough, MA), and Cobas 6800 System (Roche Diagnostics, Basel, Switzerland). Excess positive nasopharyngeal/oropharyngeal swabs were stored at -80C prior to processing for WGS.

Specimens were processed using custom NEB/Roche reagents the SARS-CoV-2 v2 SNAP kit + Omicron spike-in from IDT/Swift and were sequenced on the Illumina NovaSeq 6000 platform on SP or S2 flow cells. Variant calls were made for each specimen based on the proportion of variant-defining mutations present in the specimen (cutoff: ≥40%). Specimens that did not meet a variant-defining criterion were classified as an “unidentified variant.” Specimens with ≤65% of targeted bases covered at 100x were considered to have failed quality control and excluded. This cutoff was selected internally as it corresponded with the inclusion of 5% of unclassifiable samples (e.Table1).

### Data Access

Demographic, comorbidity, COVID-19 vaccination, SARS-CoV-2 testing, and hospitalization data were extracted from the electronic health records (HER) using the Yale Computational Health Platform.[25] Records for vaccinations that occurred outside of YNHH were obtained from the state vaccination registry and extracted using the same platform. COVID-19 symptom data were collected from medical notes using a Natural Language Processor (NLP).[26] This study was approved by the Yale Institutional Review Board (ID#2000030222).

### Study Sample

We identified all SARS-CoV-2 RT-PCRs collected among vaccine eligible people (ages ≥16 years) in the YNHH EHR between April 1 and August 24, 2021. We excluded tests from people who received a vaccine prior to state distribution (December 14, 2020), had a previous positive SARS-CoV-2 RT-PCR or rapid antigen test, or had missing covariate information (see Statistical Analysis). Additionally, we excluded tests that were performed after receiving a booster (3^rd^) dose or an Ad26.COV2 vaccine dose.

### Outcome (Case) Definition and Control Selection

Our primary outcomes of interest were WGS-classified Alpha-, Delta-, and Other-variants infection, symptomatic infection, and COVID-19 associated hospitalizations. We defined infection as positive, sequenced specimens that passed quality control. Infections were categorized as Alpha, Delta, or Other-Variant based on their WGS classification, where the Other-Variant category comprised all non-Alpha, non-Delta samples. Controls were defined as negative RT-PCRs. We selected up to three negative tests (controls) per person. If an individual had more than one negative test within a seven-day period, one random test was selected.

Symptomatic SARS-CoV-2 infection was defined as the subset of SARS-CoV-2 infections identified among symptomatic people (people with at least one NLP-captured COVID-19-related symptom recorded within 0-14 days prior to or following testing). We defined COVID-19 associated hospitalizations as the subset of symptomatic SARS-CoV-2 infections collected in the 21 days prior to or three days following hospitalization. Controls for these outcomes were selected among negative RT-PCR test results from symptomatic people.

Our secondary outcomes were calendar period-classified Alpha and Delta infections. Specifically, we defined Alpha and Delta infections as positive RT-PCR tests collected during periods when the variant comprised ≥50% of sequenced samples from Connecticut deposited in the global initiative on sharing all influenza data (GISAID; https://www.gisaid.org).[27,28] Alpha accounted for ≥50% of sequenced samples from the beginning of the study (April 1) through May 28, 2021.[27,28] Delta contributed ≥50% of the sequenced samples beginning on June 27, 2021.[27,28] Controls were then limited to negative RT-PCRs collected during the period used for variant classification.

### Statistical Analysis

We visually summarized the number of infections, or positive RT-PCRs collected in the absence of a positive test in the previous 90 days, recorded in the YNHH between April 1 and August 24, 2021, and vaccine coverage among selected cases and controls by day. The number of sequenced samples that passed quality control were visually summarized by day and variant classification.

#### Vaccine Effectiveness Against WGS-Classified SARS-CoV-2 Outcomes

We estimated the association between mRNA COVID-19 vaccination (<14 days, 14-89 days, 90-149 days, and ≥150 days since 2^nd^ dose) and WGS-classified Alpha, Delta, and Other-Variant infection, symptomatic infection, and COVID-19 associated hospitalization using generalized additive multinomial, logistic regressions. We included the following *a priori* selected covariates: date of test (continuous), age (continuous), sex, race/ethnicity, Charlson comorbidity score [29] (continuous), number of non-emergent YNHH encounters in the year prior to vaccine rollout in Connecticut (December 2020; categorized as 0, 1-2, 3-4, 5+), insurance group (uninsured, Medicaid, Medicare, other), social vulnerability index of residential zip code (continuous) and residential county. Continuous factors were modeled using a natural spline with 3 knots.[30,31] From the model we estimated vaccine effectiveness as one minus the odds ratio (OR) of infection comparing vaccinated to unvaccinated people.

#### Duration of Protection from Primary Vaccination

We tested for declines in the level of protection over time by comparing the odds of infection among recently vaccinated people (14-89 days since 2^nd^ dose) to the odds of infection among people who received their 2^nd^ dose 90-149, and ≥150 days prior to testing.[32] We evaluated this association using a generalized additive logistic regression and accounted for the same confounders as in the WGS-classified vaccine effectiveness analysis.

#### Calendar Period-Classified, Variant-Specific Vaccine Effectiveness

We estimated period defined variant specific vaccine effectiveness using generalized additive logistic regressions and included the same confounders as in the WGS-classified vaccine effectiveness analysis. All analyses were conducted in R, version 4.1.2.[33]

### Sensitivity Analyses

We performed multiple sensitivity analyses testing the robustness of our findings to alternative study design, data cleaning and modeling assumptions. Specifically, we examined the following scenarios: matched analysis (1:4 matching with replacement) and various quality control definitions. Additionally, we estimated vaccine effectiveness and tested for differences in the level of protection during periods when variants were co-circulating. Finally, to ensure that any differences observed between the period-classified and WGS-classified effectiveness estimates were not the result of bias introduced through the selection of sequenced samples, we performed the period-classified analysis among sequenced samples. For a detailed description, see *Supplement: Sensitivity Analyses*.

## Results

The regions served by YNHH experienced two successive waves of SARS-CoV-2 infections (or positive RT-PCRs) between April 1 and August 24, 2021 (Figure1.A). According to the 4,125 samples available for sequencing during this period (1,076 of which failed quality control and were omitted), the first wave was comprised of Alpha and Other-Variant (non-Alpha/Delta) infections and the second was predominantly comprised of Delta infections (Figure1.B). The proportion of tests collected among vaccinated people increased during April but remained around 50% for the rest of the study period (Figure1.C).

**Figure 1.**
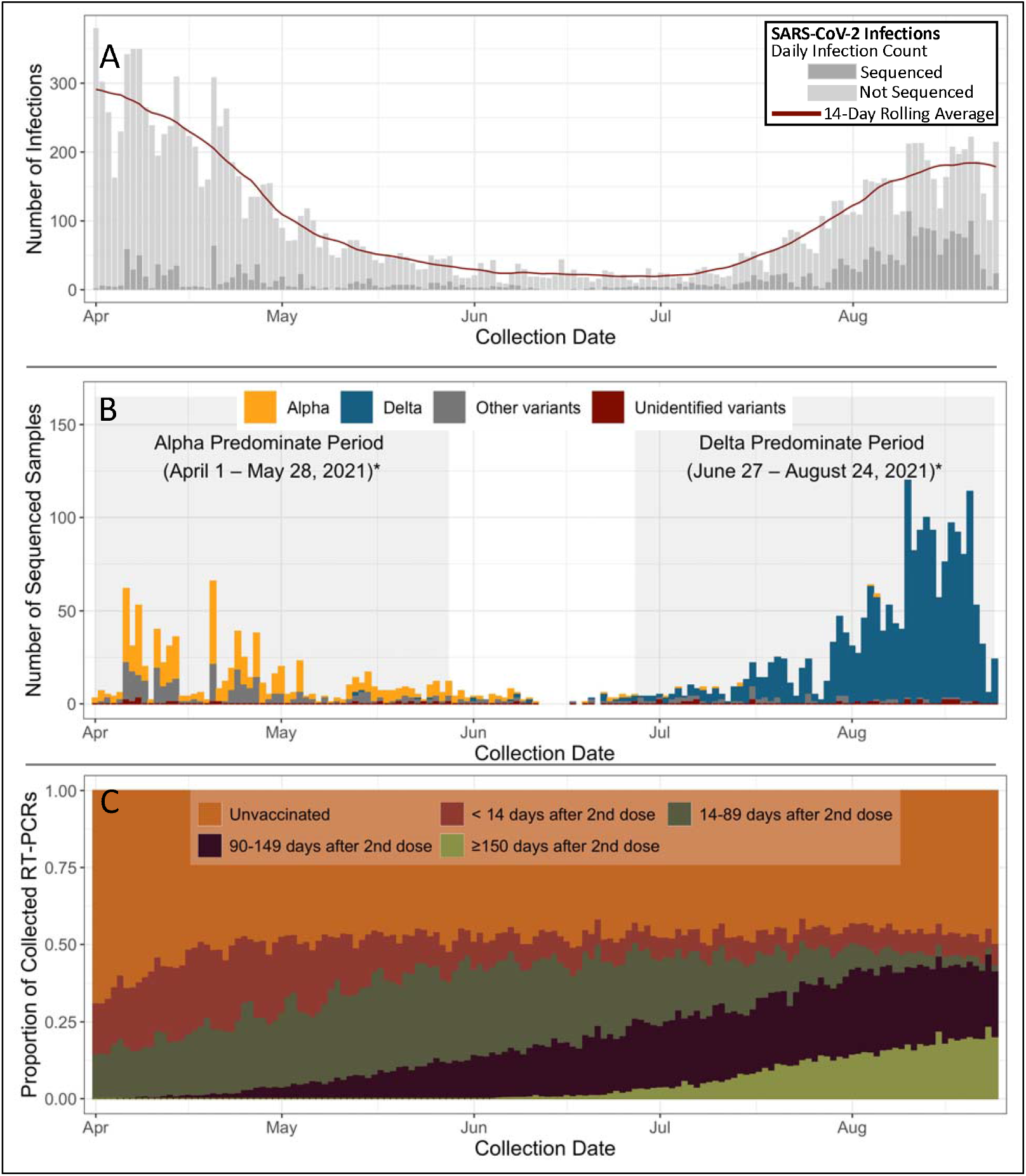
Trends in SARS-CoV-2 Infection Frequency and Vaccination Status at the Yale New Haven Health System between April 1, 2021, and August 24, 2022. The trends in (A) SARS-CoV-2 infections, (B) whole genome sequence (WGS) defined variant specific infections that passed quality control overlayed on the Alpha and Delta predominant periods (Alpha: April 1 – May 28, 2021, Delta: June 27 – August 24, 2021), and (C) mRNA vaccination status over the study period (April 1, 2021, through August 24, 2022). SARS-CoV-2 infections were defined as positive RT-PCRs collected among people without a prior positive SARS-CoV-2 RT-PCR recorded in the Yale New Haven Health System. Infections were stratified by if they were sequenced (dark grey) or not (light grey). Variant predominant periods were defined as periods where the variant comprised at least 50% of CT samples in GISAID.

### Population and Sample

Between April 1 and August 24, 2021, 502,618 RT-PCRs were collected among 268,045 vaccine eligible people. Following the restriction to RT-PCRs that met the inclusion criteria (n=441,356 tests among 241,654 people), the sample contained 10,349 positives (cases), 2,565 (25%) of which were sequenced (1,560 sequenced samples did not meet our studies inclusion criteria) and 2,029 passed quality control. From the 431,007 negative RT-PCRs that met our inclusion criteria, we randomly selected up to three per person, resulting in the inclusion of 343,727 negative RT-PCRs as controls (Figure2).

**Figure 2:**
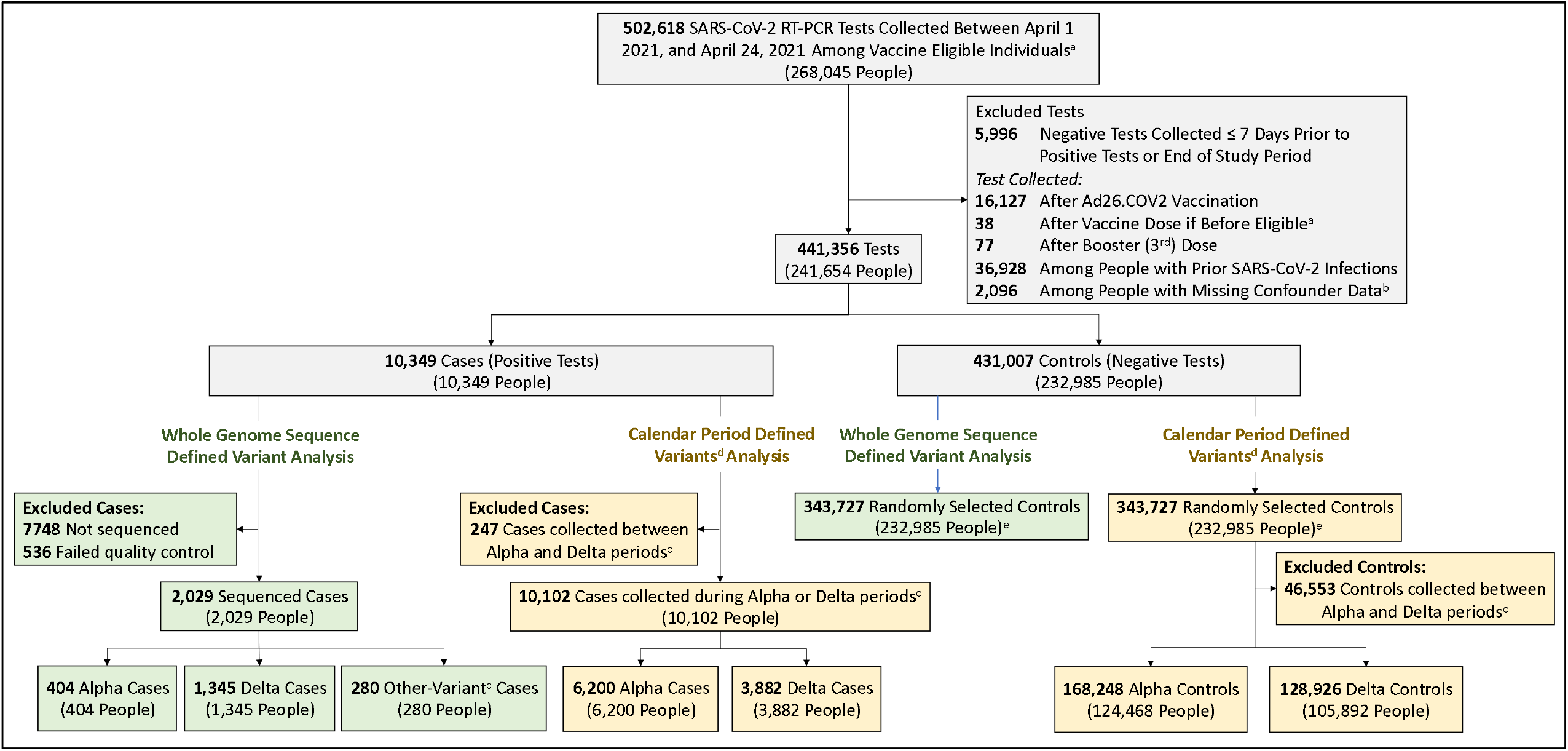
Selection of Tests for the Case Control Analysis. The sample was limited to viral RNA samples collected between April 1 and August 24, 2021, among vaccine eligible individuals (people aged 16 years or greater^a^) and had recorded confounder data (2,032 and 64 tests were collected among people without SVI and sex data, respectively). Whole genome sequencing was performed on available specimens and variant calls were made based on the frequency of variant defining mutations. The other-variant classification incorporated all non-Alpha/Delta samples^c^. Period based variant classifications were defined as cases collected during variant dominate periods, or periods when at least 50% of sequenced samples deposited in GISAID were that variant.[28] The Alpha dominate period was April 1 through May 28, 2021 and the Delta dominate period was July 10 through August 24, 2021^d^. People were allowed to contribute up to three negative tests to the control sample^e^.

Cases (positive RT-PCRs), sequenced cases and controls were similar with respect to age, gender, SVI of residential zip code and Charlson comorbidity score. However, a larger proportion of controls occurred among non-Hispanic White people (61% of controls vs 40% of Alpha infections, 49% of Delta infections, and 41% of Other-Variant infections). Among vaccinated people, the median time between 2^nd^ dose administration and testing was shorter for Alpha cases (55 days [1^st^-3^rd^ Quartile (1-3Q): 25-66 days]) than Other-Variant cases (85 days [1-3Q: 54-127 days]) and Delta cases (135 days [1-3Q: 110-169 days], Table1).

**Table 1:** Characteristics of SARS-CoV-2 RT-PCR Tests Analyzed between April 1, 2021, and August 24, 2022 and Included as Cases or Controls.

|  | Control<sup>a</sup><br>(N=343727) | Case<sup>a</sup><br>(N=10349) | Sequenced Cases |  |  |
| --- | --- | --- | --- | --- | --- |
|  |  |  | Alpha<sup>a</sup><br>(N=404) | Delta<sup>a</sup><br>(N=1345) | Other<sup>a</sup><br>(N=280) |
| Age [Median (p25-p75)] | 48 [31, 65] | 40 [28, 55] | 38 [27, 54] | 41 [29, 58] | 41 [28, 59] |
| Sex [N (%)] |  |  |  |  |  |
| Female | 201942 (58.8%) | 5651 (54.6%) | 224 (55.4%) | 695 (51.7%) | 160 (57.1%) |
| Male | 141785 (41.2%) | 4698 (45.4%) | 180 (44.6%) | 650 (48.3%) | 120 (42.9%) |
| Race Ethnicity [N (%)] |  |  |  |  |  |
| Black or African American | 40524 (11.8%) | 1854 (17.9%) | 97 (24.0%) | 249 (18.5%) | 59 (21.1%) |
| Hispanic or Latino | 44327 (12.9%) | 2316 (22.4%) | 104 (25.7%) | 264 (19.6%) | 66 (23.6%) |
| Other/Unknown | 49575 (14.4%) | 1270 (12.3%) | 43 (10.6%) | 169 (12.6%) | 39 (13.9%) |
| White | 209301 (60.9%) | 4909 (47.4%) | 160 (39.6%) | 663 (49.3%) | 116 (41.4%) |
| Charlson Score<sup>c</sup> [Median (p25-p75)] | 0 [0, 1.00] | 0 [0, 1.00] | 0 [0, 1.00] | 0 [0, 1.00] | 0 [0, 1.00] |
| SVI [Median (p25-p75)] | 0.52 [0.45, 0.52] | 0.52 [0.45, 0.52] | 0.52 [0.52, 0.52] | 0.52 [0.45, 0.52] | 0.52 [0.45, 0.52] |
| Insurance Group [N (%)] |  |  |  |  |  |
| Uninsured | 19168 (5.6%) | 858 (8.3%) | 31 (7.7%) | 130 (9.7%) | 31 (11.1%) |
| Medicaid | 49306 (14.3%) | 2559 (24.7%) | 135 (33.4%) | 279 (20.7%) | 60 (21.4%) |
| Medicare | 36640 (10.7%) | 554 (5.4%) | 23 (5.7%) | 99 (7.4%) | 13 (4.6%) |
| Other | 238613 (69.4%) | 6378 (61.6%) | 215 (53.2%) | 837 (62.2%) | 176 (62.9%) |
| Known of Non-Emergent Visits<sup>d</sup> [Median (p25-p75)] | 1.00 [0, 8.00] | 0 [0, 5.00] | 1.00 [0, 8.00] | 0 [0, 5.00] | 1.00 [0, 7.00] |
| Vaccination Status at time of test [N (%)] |  |  |  |  |  |
| Unvaccinated | 171149 (49.8%) | 7966 (77.0%) | 340 (84.2%) | 873 (64.9%) | 223 (79.6%) |
| Incomplete primary vaccination (<14 days after 2<sup>nd</sup> dose) | 47505 (13.8%) | 988 (9.5%) | 41 (10.1%) | 59 (4.4%) | 28 (10.0%) |
| Complete primary vaccination |  |  |  |  |  |
| 14-89 Days after 2<sup>nd</sup> dose | 63296 (18.4%) | 369 (3.6%) | 22 (5.4%) | 45 (3.3%) | 14 (5.0%) |
| 90-149 Days after 2<sup>nd</sup> dose | 44475 (12.9%) | 633 (6.1%) | 1 (0.2%) | 217 (16.1%) | 10 (3.6%) |
| ≥150 Days after 2<sup>nd</sup> dose | 17302 (5.0%) | 393 (3.8%) |  | 151 (11.2%) | 5 (1.8%) |
| Interval between 2<sup>nd</sup> dose and testing [Median (p25-p75)] | 83 [45, 121] | 115 [78, 152] | 55 [25, 66] | 135 [110, 169] | 85 [54, 127] |

<sup>a</sup> Participants allowed to contribute both cases (positive RT-PCRs) and up to three controls (negative RT-PCRs) tests
<sup>b</sup> SARS-CoV-2 variants defined based on whole genome sequencing
<sup>c</sup> Score as of December 2020
<sup>d</sup> Number of non-emergent visits in the year prior to the vaccination period within YNHH (December 2nd 2019-December 1st 2020)

### Vaccine Effectiveness Against WGS-classified SARS-CoV-2 Outcomes

The effectiveness of primary mRNA vaccination 14-89 days after 2^nd^ dose administration was 84.4% (95% Confidence Interval [CI]: 75.6-90.0%) and 68.9% (CI: 58.0-77.1%) against Alpha and Delta infection, respectively, and 85.5% (CI: 65.5-93.9%) and 89.0% (CI: 52.9-97.4%) against Alpha and Delta associated hospitalizations, respectively (Figure3.A/C). The effectiveness of primary vaccination did not differ between Alpha-, Delta-, and Other-Variant classified infections for people who received their second dose <14 days prior to testing. However, the level of protection offered against Alpha infection was significantly higher than that offered against Delta infection for people who received their second dose 14-89 days prior to testing (Pvalue: 0.013). Only one Alpha infection was recorded among people vaccinated ≥90 days prior to testing and we were unable to reliably compare the level of protection ≥90 days after second dose receipt. The level of protection offered against Other-Variant infection was lower than against Alpha infection and higher than Delta infection but was not significantly different from either. The level of protection did not vary significantly by variant for symptomatic infection or COVID-19 associated hospitalization (Figure2).

**Figure 3:**
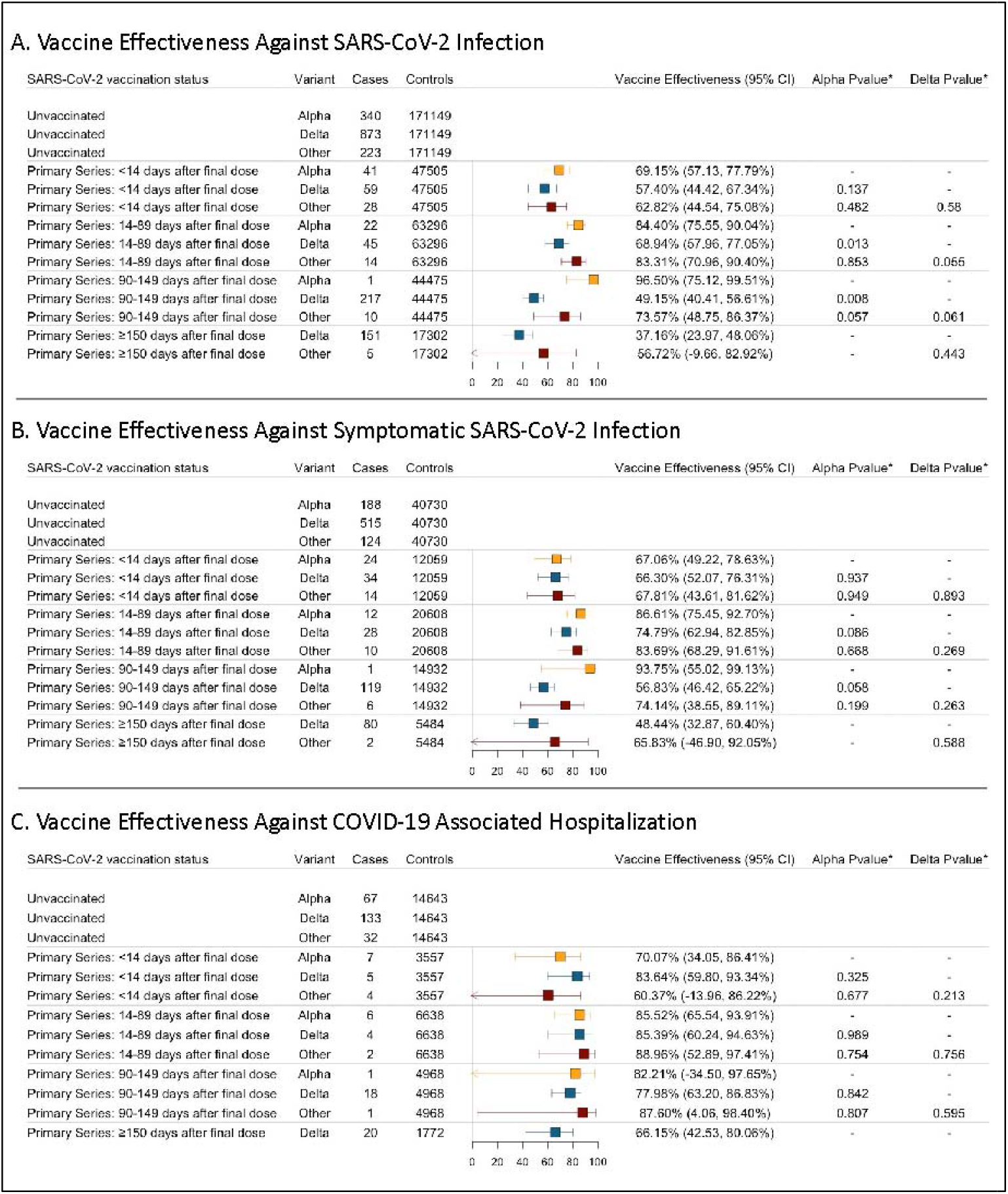
Forest Plot of mRNA Vaccine Effectiveness Against Whole Genome Sequence Classified Alpha, Delta, and Other-Variant SARS-CoV-2 Infection, Symptomatic Infection, and COVID-19 Associated Hospitalization Identified at the Yale New Haven Hospital System between April 1, 2021, and August 24, 2022. The effectiveness of primary mRNA vaccination against variant defined (A) SARS-CoV-2 infection, (B) symptomatic infection, and (C) COVID-19 associated hospitalization. (A) SARS-CoV-2 infection was defined as a RT-PCR positive, sequenced viral RNA samples collected among a vaccine eligible person (≥16 years old at testing) and passed quality control (≥65% bases covered at 100x). (B) Symptomatic infection was defined as a SARS-CoV-2 infection collected within 12 days of a record of COVID-19 symptoms (captured using a natural language processor). (C) COVID-19 associated hospitalization was defined as a symptomatic infection that was collected within the three days after to or 21 prior to hospitalization. Cases were defined as Alpha, Delta, and Other-Variant based on whole genome sequencing. The other-variant classification incorporated all non-Alpha/Delta infections. Rows containing no cases were omitted. *The levels of protection offered against the examined variants were compared and significance was defined with an alpha of 0.05. (RT-PCR: reverse transcription polymerase chain reaction)

### Duration of Protection from Primary Vaccination

The odds of Delta infection and symptomatic infection were significantly higher 90-149 and ≥150 days after 2^nd^ dose administration than 14-89 days after administration (e.Table2). However, the odds of Delta associated hospitalization, as well as Alpha and Other-Variant infection, symptomatic infection, or COVID-19 associated hospitalization, did not increase significantly. The precision around these estimates was, however, low.

### Calendar Period-Classified, Variant-Specific Vaccine Effectiveness

A total of 6,200 cases and 168,248 controls were collected during the Alpha predominant period (April 1-May 28, 2021). During the Delta predominant period (June 27-August 24, 2021), 3,882 cases and 128,926 controls were collected (Figure2). Cases collected during the variant predominant periods were similar to sequenced cases with respect to demographic and clinical factors. However, the median time between second dose receipt and calendar-period classified Alpha infections was shorter (37 days) than WGS-classified Alpha infections (55 days, eTable3).

Among the samples collected during the Alpha predominant period, 65% were Alpha, 0.9% were Delta, and 34.5% were Other-Variant. Conversely during the Delta period, 96% of sequenced samples were Delta (eTable4). The effectiveness of primary vaccination 14-89 days after 2^nd^ dose administration was 88.2% (95% CI: 86.3-89.8%) and 64.6% (95% CI: 58.0-70.4%) during the period of Alpha and Delta predominance, respectively (Figure4).

**Figure 4:**
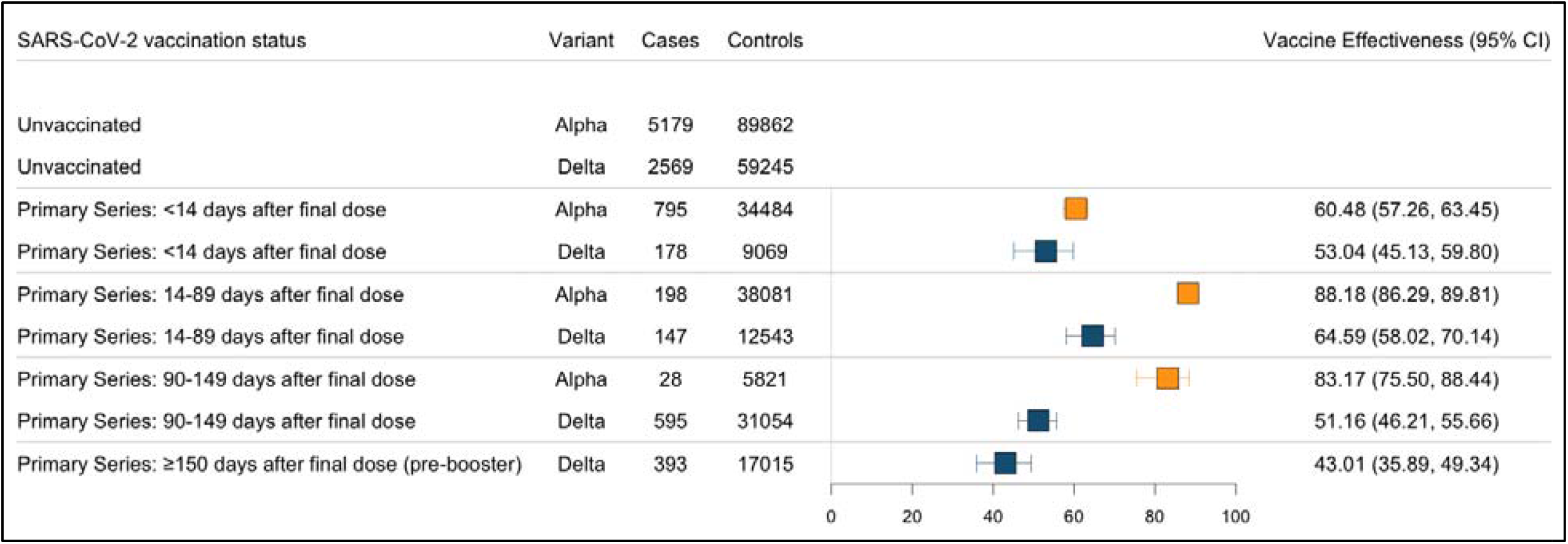
Forest Plot of mRNA Vaccine Effectiveness Against Calendar-Period Classified Alpha, Delta, and Other-Variant SARS-CoV-2 Infection Identified at the Yale New Haven Hospital System between April 1, 2021, and August 24, 2022. The effectiveness of primary mRNA vaccination against variant defined SARS-CoV-2 infection. Infection was defined as a RT-PCR positive samples collected among a vaccine eligible person (≥16 years old at testing). Infections were classified based on periods of variant predominance (≥50% of Connecticut samples deposited in GISAID). The Alpha dominate period was April 1 through May 28, 2021 and the Delta dominate period was July 10 through August 24, 2021.

### Sensitivity Analyses

The effectiveness of primary vaccination (14-89 days after 2^nd^ dose administration) ranged between 81.5-89.9% against Alpha and 67.3-70.4% against Delta in the sensitivity analyses (eFigure1-7/eTable5). Under each examined scenario, the level of protection ≥90 days after 2^nd^ dose administration was significantly higher against Alpha than against Delta. The effectiveness of vaccination ≥14 days after 2^nd^ dose administration was significantly higher against Alpha than Delta for all examined scenarios except when quality control was defined at 100% bases covered at 100x (eFigure2). The level of protection offered against Other-Variant infection was higher than that offered against Delta infection for the matched and quality control defined as 50% bases covered at 100x analyses (e.Figure1/3). The effectiveness against calendar-period classified infections were similar following the restriction to sequenced samples (14-89 days Alpha: 84.4% [95% CI: 77.1-89.4%]; Delta: 69.3% [95% CI: 58.6-77.2%], e.Table5) as the primary analysis (14-89 days Alpha: 88.2% [95% CI: 86.3-89.8]; Delta: 64.6% [95% CI: 58.0-70.1%], Table2).

## Discussion

In this retrospective analysis, we estimated the effectiveness of primary mRNA COVID-19 vaccination against WGS-classified, variant-specific SARS-CoV-2 infection, symptomatic infection, and COVID-19 associated hospitalization. We found that mRNA COVID-19 vaccines provided less protection against Delta infection than Alpha infection and that the level of protection offered against Delta infection declined significantly over time since vaccine administration. However, we did not observe a significant difference in the level of protection offered against variant-specific symptomatic infection or hospitalization.

These findings build upon existing literature suggesting that the increase in SARS-CoV-2 cases among vaccinated people during the Delta wave of fall 2021 was the result of variant-specific immune evasion and waning levels of vaccine protection.[3–5,10–17,34–38] However, while there is ample evidence of both phenomena during the Delta period [3–5,10–17,34–38], few studies have disentangled the two processes.[3–5,10–17,34–38] Further, even when their cooccurrence was reported, the authors have typically concluded that one of the two phenomena was the primary driver of the reduced effectiveness.[13,14,16] In contrast, we, in alignment with Britton et al., observed strong evidence for both declining levels of protection and variant-specific immune evasion.[18]

While many factors likely contribute to the variability of vaccine effectiveness estimates in the literature, differences in variant classification methodology are a potential component.[39] To examine how the vaccine effectiveness estimates from calendar-period-based variant classifications compares to WGS, we estimated the effectiveness of primary vaccination against Alpha and Delta cases defined by periods of variant predominance. We found that estimates of vaccine effectiveness against SARS-CoV-2 infection during periods of Alpha and Delta predominant were similar to those estimated using WGS.

These findings are not surprising. While there was a high degree (35% of sequenced samples) of misclassification during the Alpha predominance period, effectiveness against Alpha and other-variant (the major variant category co-circulating during the Alpha predominant period) infection did not vary significantly in the WGS analysis. Thus, we would not expect that this degree of misclassification would significantly alter vaccine effectiveness estimates. However, this finding is unlikely to hold under situations with similarly large amounts of misclassification and underlying differences in variant-specific vaccine effectiveness.

Unlike Alpha, data from the WGS analysis suggests that the effectiveness against Delta infection compared to other-variant infections may differ (differed significantly for multiple sensitivity analyses but not primary analysis). As a result, a large amount of variant misclassification during the Delta predominant period may impact vaccine effectiveness estimates. However, Delta rapidly became the predominant variant accounting for 50% of samples from Connecticut in GISAID on June 27, 2021, 75% on July 10, 2021, and 100% on September 6, 2021.[27,28] It is, thus, unsurprising that the period-classified vaccine effectiveness estimates mirrored the WGS-classified estimates.

These findings suggest that, even when the effectiveness of the variants is expected to differ, calendar-period-based variant classification may yield reliable estimates, when there is limited superimposition of variant waves, and the proportion of misclassified cases is expected to be low. However, under scenarios when variants or subvariants are co-circulating, as is the case with the Omicron subvariants, calendar-period-based classification provides limited utility. Not only would subvariant calendar-period-based classification result is meaningful amounts of misclassification, but it also prevents researchers from comparing the effectiveness of variants as they co-circulate. As a result, the need for WGS in future vaccine effectiveness and severity analysis has only increased.

### Limitations

Our study was subject to several limitations. First, specimens available for sequencing were limited. However, the storage and sequencing of specimens was considered random and, while it reduced our power, this restriction was unlikely to have introduced bias. Further, our sensitivity analysis limiting the calendar-period classified infections to sequenced samples mirrored the main analysis. Second, to classify variants using WGS we had to exclude cases that did not meet our quality control definition. While this further reduced our sample, we show that this restriction was unlikely the driver of our results in sensitivity analyses. Third, sequenced samples have lower CT values than those for which sequencing is unsuccessful. For this reason, the vaccine effectiveness estimates against infection may be biased towards more severe illness, which in turn may impact the generalizability of our findings. Finally, we had few to no Alpha outcomes for the ≥90 days after 2^nd^ dose administration categories and were unable to provide reliable estimates.

## Conclusion

In this study, the use of WGS revealed differences in variant-specific COVID-19 vaccine effectiveness, with the effectiveness of primary mRNA vaccination against Delta infection being significantly lower than that against Alpha infection. However, the effectiveness of vaccination was found to be moderate against symptomatic COVID and high against COVID-19 associated hospitalization regardless of variant. Although we observed broad agreement between estimates of variant-specific vaccine effectiveness when WGS and calendar-period were used to define Alpha and Delta variants, there was a significant degree of misclassification associated with calendar-period classifications, which may limit its application in future SARS-CoV-2 epidemic waves and in settings with heterogeneity in variant-specific vaccine effectiveness.

## Data Availability

Data cannot be shared publicly because of the presence of potentially identifiable health information. Requests for access can be directed to the corresponding author and Yale Human Research Protection Program.

## Author Contributions

Margaret L. Lind, Albert I. Ko and Wade L. Schulz have full access to all the data in the study and take responsibility for the integrity of the data and the accuracy of the data analysis.

*Concept and design:* Lind, Hitchings, Schulz, Ko, Cummings, Hamilton, McCarthy, Copin

*Acquisition and interpretation of data:* Lind, Warner, Coppi, Duckwall, Ferguson, Coppi

*Drafting of the manuscript:* Lind, Hitchings, McCarthy, Copin, Ko

*Critical revision of the results:* All authors.

*Critical revision of the manuscript for important intellectual content:* All authors.

*Statistical analysis:* Lind

*Administrative, technical, or material support:* Duckwall, Borg

*Supervision:* Ko, Hitchings, Schulz, Cummings, Hamilton

## Supplement

**eTable1:** Variant call distribution by Percent Bases Covered (100x)

**eTable2:** Risk of Variant Specific SARS-CoV-2 Infection among Vaccinated People*, According to Time after Receiving a Second Primary Vaccine Dose

**eTable3:** Misclassified Alpha and Delta SARS-CoV-2 Cases Define using Calendar Period

### Sensitivity Analyses

**eFigure1:** *Forest Plot of Vaccine Effectiveness (Matching)*

**eFigure2:** *Forest Plot of Vaccine Effectiveness (Quality Control – 100% bases covered at 100x)*

**eFigure3:** *Forest Plot of Vaccine Effectiveness (Quality Control – 50% bases covered at 100x)*

**eFigure4:** Number of Sequenced Samples by Variant 21signation and Date (Daily Count and 5-Day Rolling Average)

**eFigure5:** *Forest Plot of Vaccine Effectiveness* (*Alpha and Delta Cocirculating period: May 13 – August 5, 2021)*

**eFigure6:** *Forest Plot of Vaccine Effectiveness* (*Alpha and Other Cocirculating period: April 2 – August 5, 2021)*

**eFigure7:** *Forest Plot of Vaccine Effectiveness (Delta and Other Cocirculating period: May 13 – August 21, 2021)*

**eTable 4:** Effectiveness of Primary Vaccination Against Calendar Period Defined SARS-CoV-2 Variant Infections (Sequenced Only Cases)

**eTable 1:** Variant call distribution by Percent Bases Covered (100x)

| **Percent Bases Covered** | **Samples with an identified variant call included in sample** |  | **Samples without a variant call included in sample** |  |
| --- | --- | --- | --- | --- |
|  | **Count**<br>**(N = 3230)** | **Percent of all VUS samples** | **Count**<br>**(N = 895)** | **Percent of all non-VUS samples** |
| 50% | 3063 | 94.80% | 79 | 8.80% |
| 55% | 3054 | 94.60% | 68 | 7.60% |
| 60% | 3039 | 94.10% | 56 | 6.30% |
| 65% | 3028 | 93.70% | 45 | 5.00% |
| 70% | 3017 | 93.40% | 33 | 3.70% |
| 75% | 2996 | 92.80% | 29 | 3.20% |
| 80% | 2973 | 92.00% | 23 | 2.60% |
| 85% | 2935 | 90.90% | 18 | 2.00% |
| 90% | 2880 | 89.20% | 12 | 1.30% |
| 95% | 2767 | 85.70% | 9 | 1.00% |
| 100% | 2151 | 66.60% | 7 | 0.80% |

**eTable 2:**
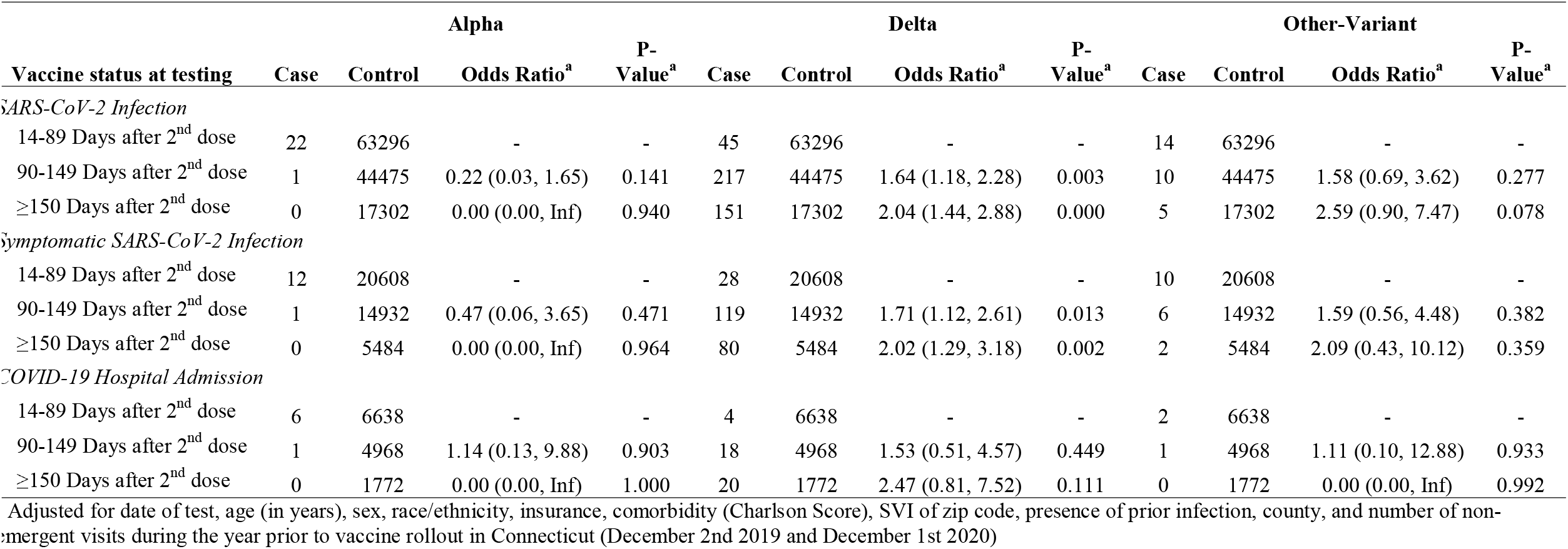
Risk of Variant Specific SARS-CoV-2 Infection among Vaccinated People*, According to Time after Receiving a Second Primary Vaccine Dose.

**eTable 3:**
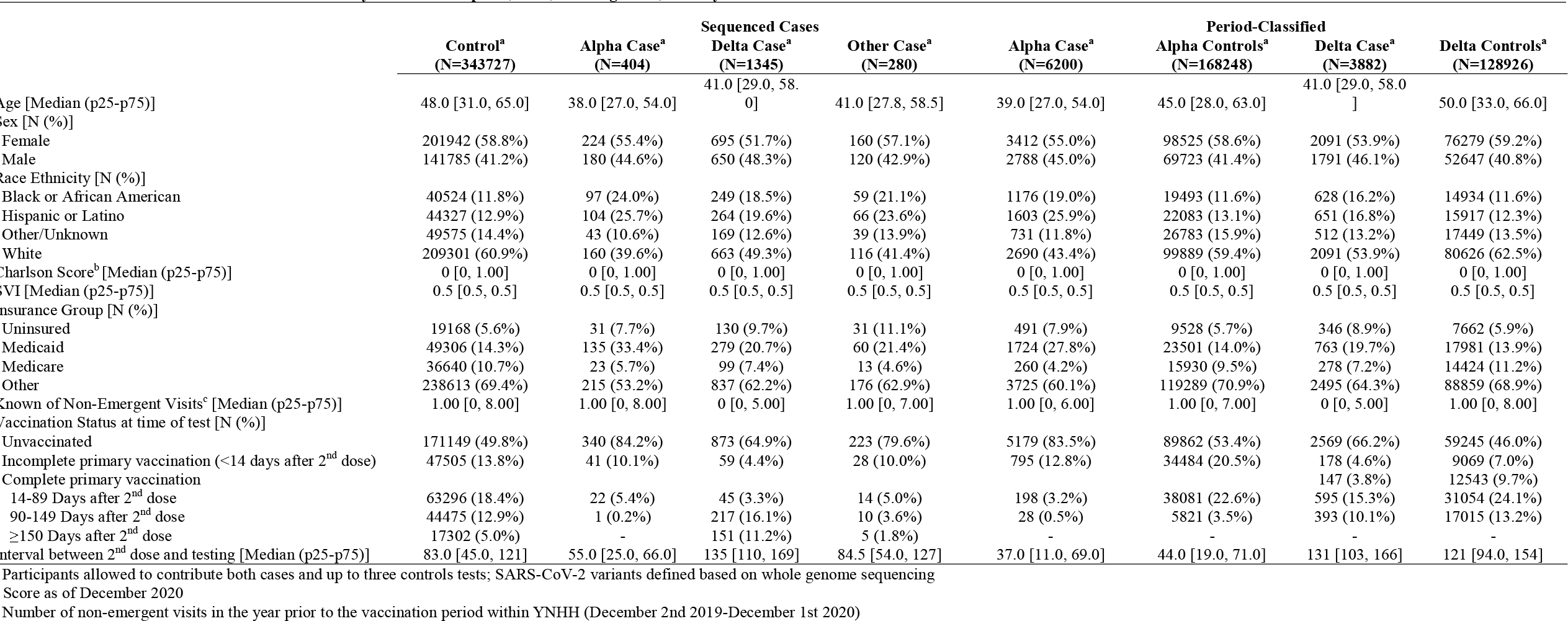
Characteristics of SARS-CoV-2 Tests Analyzed between April 1, 2021, and August 24, 2022 by Classification.

**eTable 4.** Misclassified Alpha and Delta SARS-CoV-2 Cases Define using Calendar Period.

| Period Defined Variants | Whole Genome Sequence Defined Cases |  |  |  |  |  |  |
| --- | --- | --- | --- | --- | --- | --- | --- |
|  | All Cases<br>Number | Alpha |  | Delta |  | Other Variant |  |
|  |  | Number | Percent | Number | Percent | Number | Percent |
| *Variant Call Based on Sequenced Samples Deposited in GISAID* |  |  |  |  |  |  |  |
| Alpha |  |  |  |  |  |  |  |
| ≥50% Circulating Variants<sup>a</sup> | 564 | 366 | 64.9% | 5 | 0.9% | 193 | 34.2% |
| Delta |  |  |  |  |  |  |  |
| ≥50% Circulating Variants<sup>a</sup> | 1403 | 7 | 0.5% | 1326 | 94.5% | 70 | 5.0% |

<sup>a</sup> Limited to calendar time when the variant (Alpha or Delta) accounted for a minimum of the stated percentage according to the sequenced samples deposited in the GISAID database (Alpha periods: 50% through May 28, 2021; Delta periods: ≥50% starting on: June 27, 2021, ≥75% starting on: July 10, 2021)

## Sensitivity Analyses

We tested the robustness of our findings to multiple alternative data cleaning and study designs. Under each examined scenario, we estimated vaccine effectiveness against variant specific SARS-CoV-2 infection. Each analysis mirrored the primary analyses beyond the stated changes.

### Matching (1:4 with replacement)

To allow for a single analytic sample from which to perform our analyses, we did not perform a match for our primary analysis. However, our fully adjusted (un-matched) model may suffer from positivity violations. To test if matching resulted in increased precision, we performed a 1:4 match with replacement on date of test (+/- 7 day), age group, and county. All cases successfully matched to controls.

**e.Figure 1:**
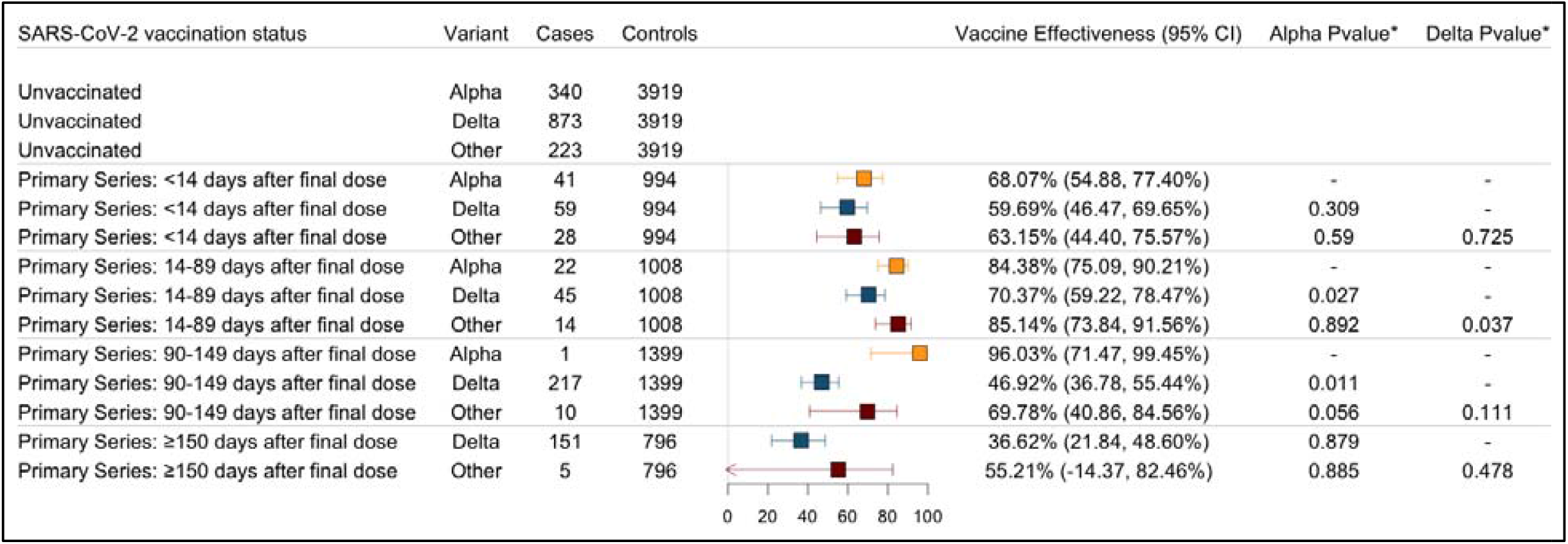
Forest Plot of Vaccine Effectiveness (Matching)

### Varied quality control definition

In the primary analysis we limited to samples that passed a quality control definition of >=65% bases covered at 100x. This quality control definition was selected as it correlated with an inclusion of a 5% potential error rate, or 5% of the included samples not having a variant call. Here we present the results from alternative quality control definitions:

- 100% bases covered at 100x
- 50% bases covered at 100x

At a threshold for inclusion of 100% bases covered (at 100x), seven sequenced samples are included without a variant designation (0.8% potential error rate) (see eTable1)

**eFigure2:**
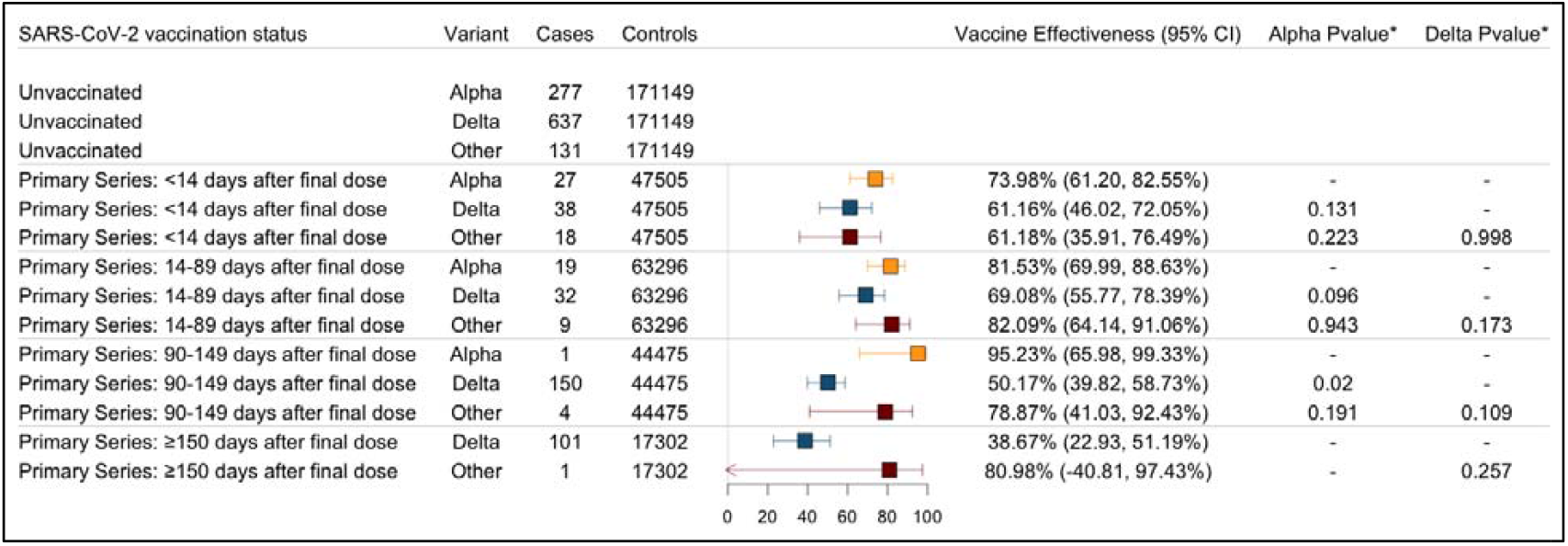
Forest Plot of Vaccine Effectiveness (Quality Control – 100% bases covered at 100x)

At a threshold for inclusion of 50% bases covered (at 100x), 79 sequenced samples are included without a variant designation (8.8% potential error rate).

**eFigure3:**
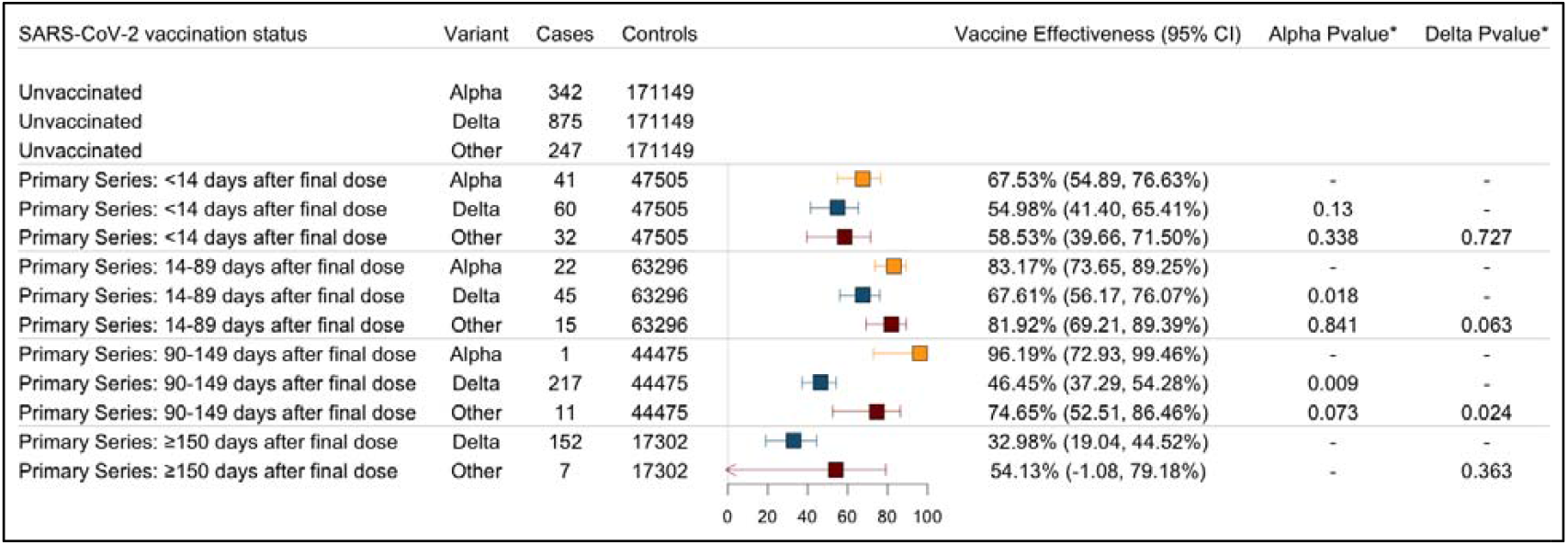
Forest Plot of Vaccine Effectiveness (Quality Control – 50% bases covered at 100x)

### Co-circulation time restricted vaccine effectiveness

In the primary analysis, we included all samples and study time into a single model and modeled calendar time flexibly. While this allowed for maximum power, the variants groups did not co-circulate the full time and our results may be impacted unaccountable, time-dependent, factors (such as changes in testing). Here, we compare the level of vaccine offered protection during periods of known variant co-circulation. We defined known circulation as the period for which we had both variants present in our included sample (based on first and last observed variant).

- Alpha and Delta: May 13 – August 5, 2021
- Alpha and Other: April 2 – August 5, 2021
- Delta and Other: May 13 – August 21, 2021

**eFigure 4:**
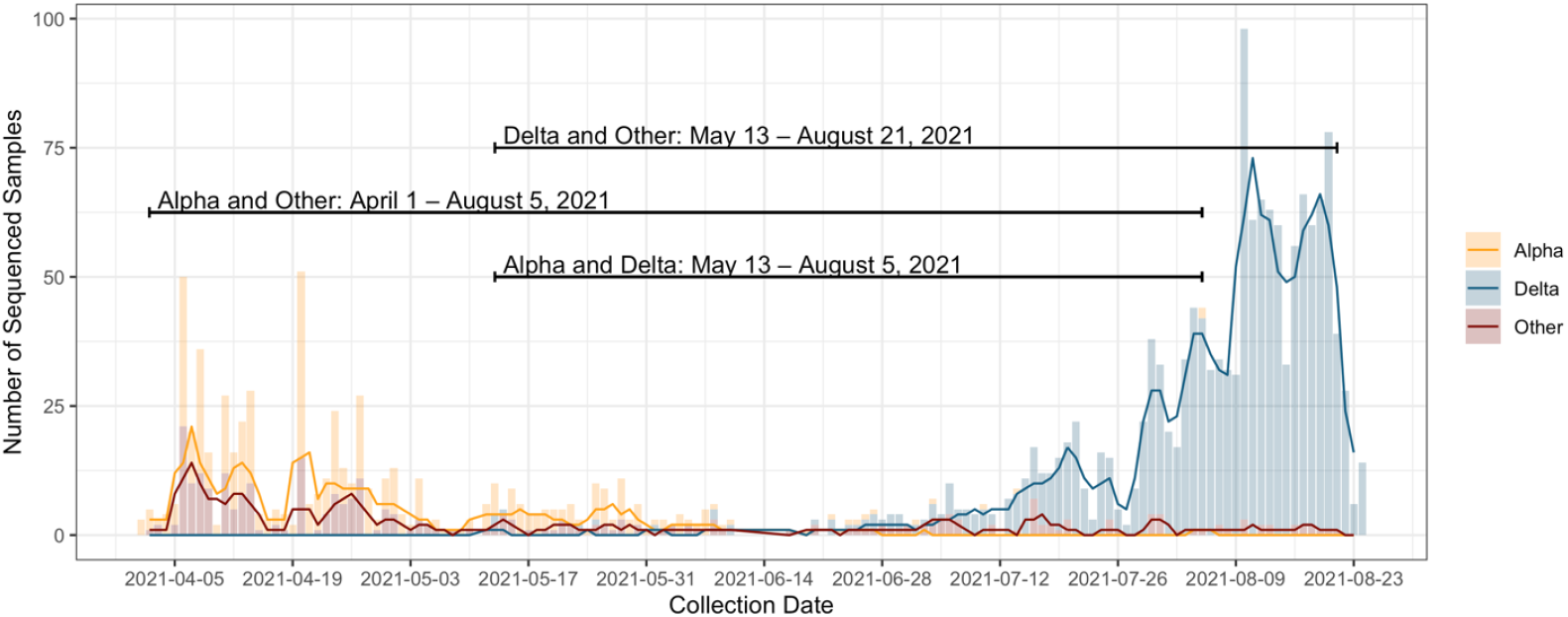
Number of Sequenced Samples by Variant 21signation and Date (Daily Count and 5-Day Rolling Average)

**eFigure5:**
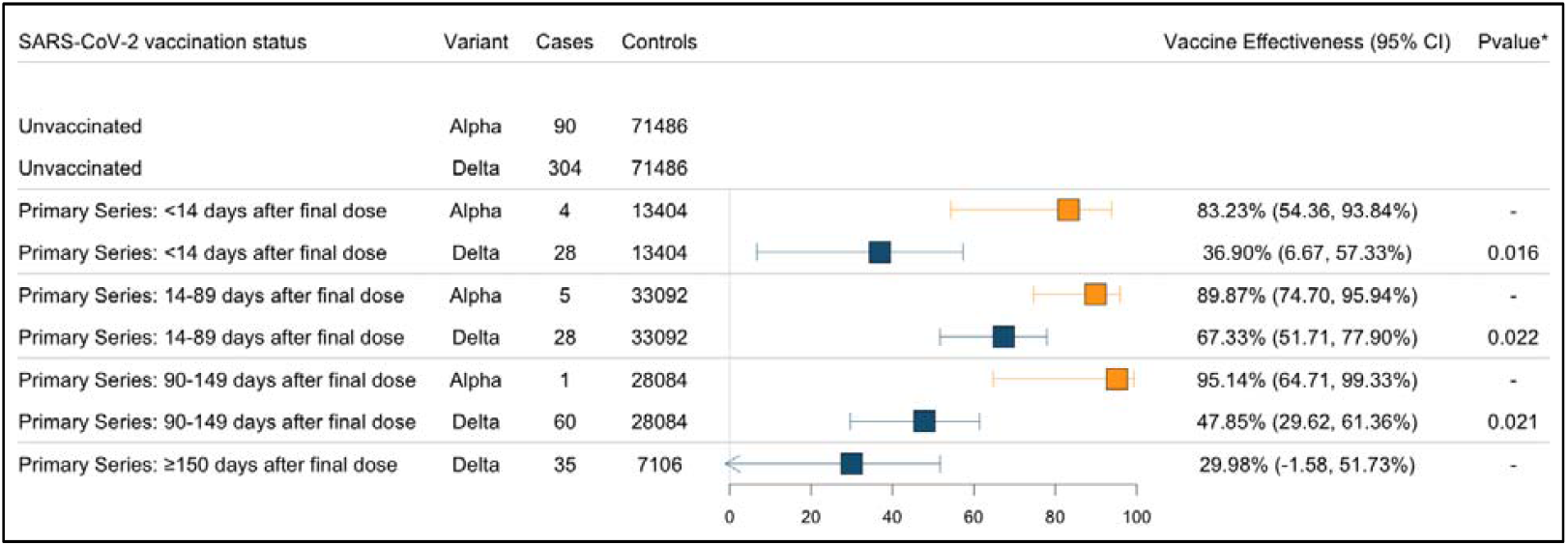
Forest Plot of Vaccine Effectiveness (Alpha and Delta Cocirculating period: May 13 – August 5, 2021)

**eFigure6:**
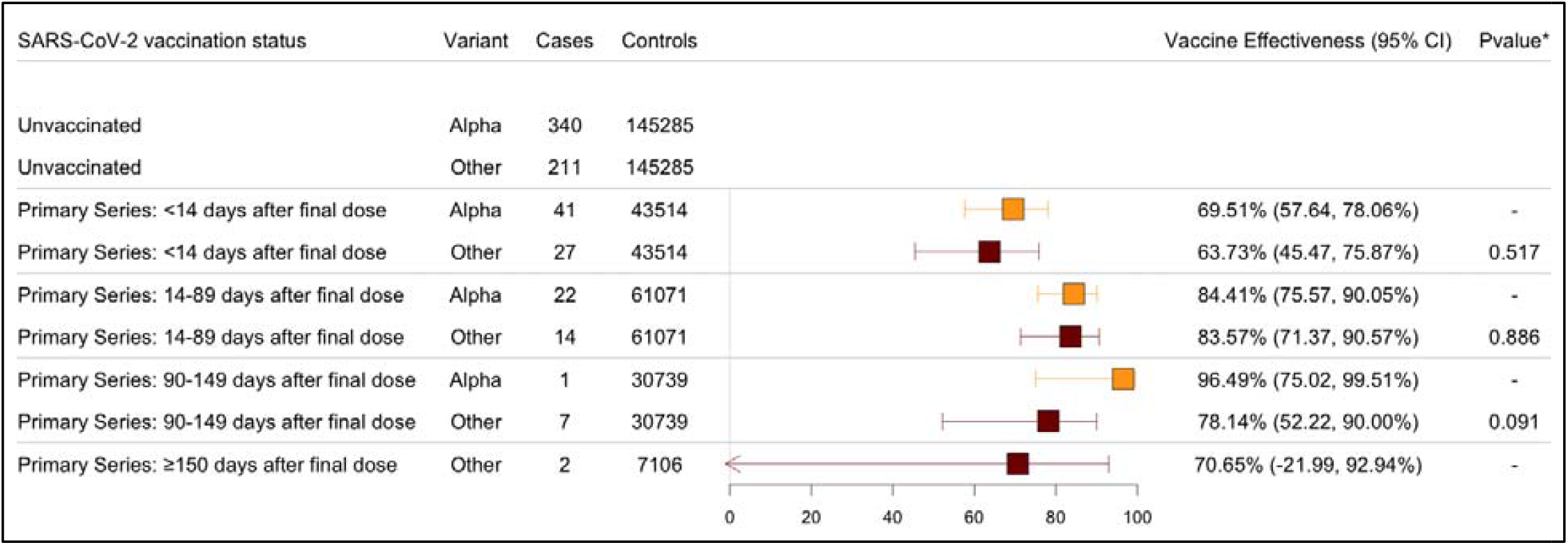
Forest Plot of Vaccine Effectiveness (Alpha and Other Cocirculating period: April 2 – August 5, 2021)

**eFigure7:**
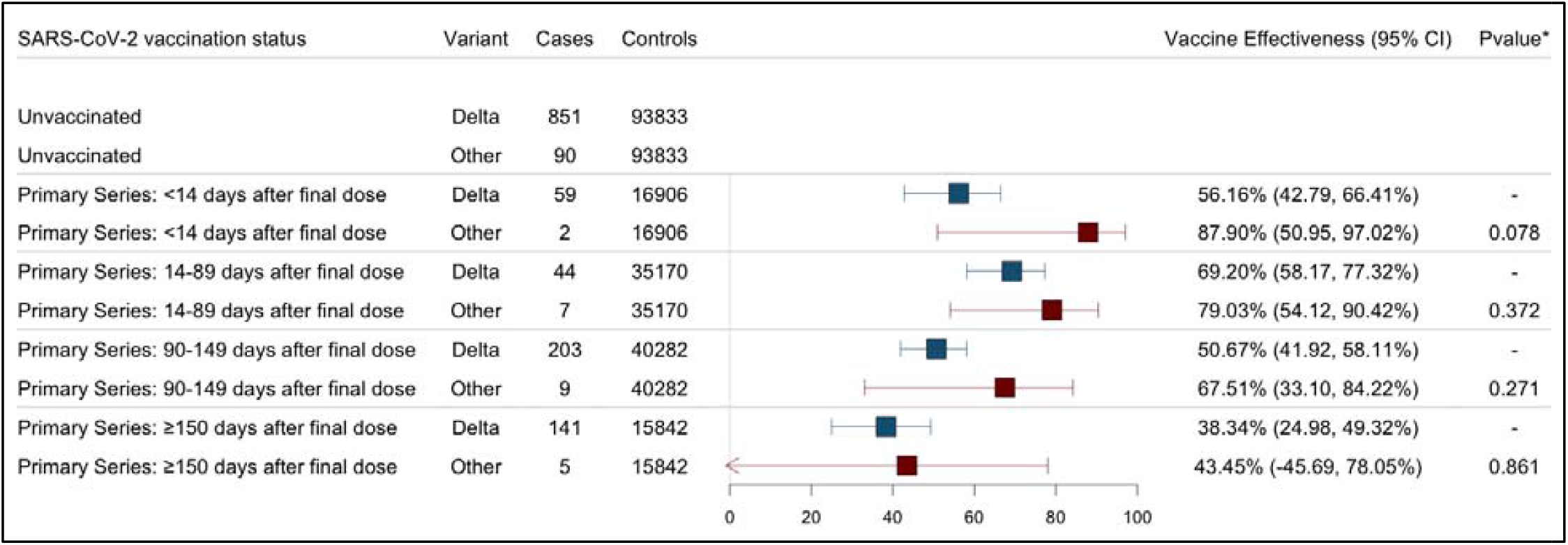
Forest Plot of Vaccine Effectiveness (Delta and Other Cocirculating period: May 13 – August 21, 2021)

### Period defined variants limited to sequenced samples

In the primary analysis, we estimated variant specific vaccine effectiveness using whole genome sequencing (WGS) and period designations. For the latter, we included all positive RT-PCRs that met our inclusion criteria (regardless of if they were sequenced). Because bias may have been introduced by the selection of specimens available for sequencing for the WGS analysis (primary), any differences we observed between these variant designation approaches may be the result of that bias. Here, we restrict the period defined analysis to sequenced samples.

**eTable 5:**
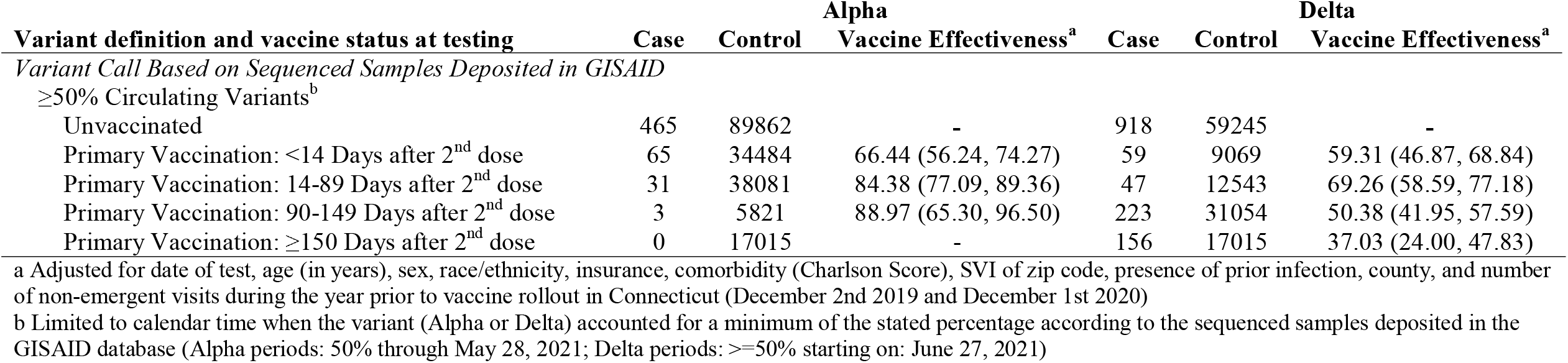
Effectiveness of Primary Vaccination Against Calendar Period Defined SARS-CoV-2 Variant Infections (Sequenced Only Cases)

## References

1. CDC. Trends in Number of COVID-19 Vaccinations in the US. 2020. Available at: https://covid.cdc.gov/covid-data-tracker. Accessed 14 June 2022.

2. CDC. Trends in Number of COVID-19 Cases and Deaths in the US Reported to CDC, by State/Territory. 2020. Available at: https://covid.cdc.gov/covid-data-tracker. Accessed 14 June 2022.

3. Puranik A, Lenehan PJ, Silvert E, et al. Comparison of two highly-effective mRNA vaccines for COVID-19 during periods of Alpha and Delta variant prevalence. medRxiv 2021; :2021.08.06.21261707.

4. Abu-Raddad LJ, Chemaitelly H, Butt AA. Effectiveness of the BNT162b2 Covid-19 Vaccine against the B.1.1.7 and B.1.351 Variants. N Engl J Med 2021; 385:187–189.

5. Bernal JL, Andrews N, Gower C, et al. Effectiveness of Covid-19 Vaccines against the B.1.617.2 (Delta) Variant. N Engl J Med 2021; Available at: https://www.nejm.org/doi/10.1056/NEJMoa2108891. Accessed 29 September 2021.

6. Eick-Cost AA, Ying S, Wells N. Effectiveness of mRNA-1273, BNT162b2, and JNJ-78436735 COVID-19 Vaccines Among US Military Personnel Before and During the Predominance of the Delta Variant. JAMA Netw Open 2022; 5:e228071.

7. Kislaya I, Rodrigues EF, Borges V, et al. Comparative Effectiveness of Coronavirus Vaccine in Preventing Breakthrough Infections among Vaccinated Persons Infected with Delta and Alpha Variants - Volume 28, Number 2—February 2022 - Emerging Infectious Diseases journal - CDC. Available at: https://www.nc.cdc.gov/eid/article/28/2/21-1789_article. Accessed 24 June 2022.

8. Rosenberg E, Dorabwila V, Easton D, et al. COVID-19 Vaccine Effectiveness by Product and Timing in New York State. Infectious Diseases (except HIV/AIDS), 2021. Available at: http://medrxiv.org/lookup/doi/10.1101/2021.10.08.21264595. Accessed 11 October 2021.

9. Tang P, Hasan MR, Chemaitelly H, et al. BNT162b2 and mRNA-1273 COVID-19 vaccine effectiveness against the SARS-CoV-2 Delta variant in Qatar. Nat Med 2021; :1–8.

10. Fowlkes A. Effectiveness of COVID-19 Vaccines in Preventing SARS-CoV-2 Infection Among Frontline Workers Before and During B.1.617.2 (Delta) Variant Predominance — Eight U.S. Locations, December 2020–August 2021. MMWR Morb Mortal Wkly Rep 2021; 70. Available at: https://www.cdc.gov/mmwr/volumes/70/wr/mm7034e4.htm. Accessed 7 October 2021.

11. Nanduri S. Effectiveness of Pfizer-BioNTech and Moderna Vaccines in Preventing SARS-CoV-2 Infection Among Nursing Home Residents Before and During Widespread Circulation of the SARS-CoV-2 B.1.617.2 (Delta) Variant — National Healthcare Safety Network, March 1–August 1, 2021. MMWR Morb Mortal Wkly Rep 2021; 70. Available at: https://www.cdc.gov/mmwr/volumes/70/wr/mm7034e3.htm. Accessed 22 June 2022.

12. Feikin DR, Higdon MM, Abu-Raddad LJ, et al. Duration of effectiveness of vaccines against SARS-CoV-2 infection and COVID-19 disease: results of a systematic review and meta-regression. The Lancet 2022; 399:924–944.

13. Bruxvoort KJ, Sy LS, Qian L, et al. Effectiveness of mRNA-1273 against delta, mu, and other emerging variants of SARS-CoV-2: test negative case-control study. BMJ 2021; 375:e068848.

14. Tartof SY, Slezak JM, Fischer H, et al. Effectiveness of mRNA BNT162b2 COVID-19 vaccine up to 6 months in a large integrated health system in the USA: a retrospective cohort study. The Lancet 2021; 398:1407–1416.

15. Pouwels KB, Pritchard E, Matthews PC, et al. Effect of Delta variant on viral burden and vaccine effectiveness against new SARS-CoV-2 infections in the UK. Nat Med 2021; 27:2127–2135.

16. Andrews N, Tessier E, Stowe J, et al. Duration of Protection against Mild and Severe Disease by Covid-19 Vaccines. N Engl J Med 2022; 386:340–350.

17. Goldberg Y, Mandel M, Bar-On YM, et al. Waning Immunity after the BNT162b2 Vaccine in Israel. N Engl J Med 2021; Available at: https://www.nejm.org/doi/10.1056/NEJMoa2114228. Accessed 29 October 2021.

18. Britton A, Fleming-Dutra KE, Shang N, et al. Association of COVID-19 Vaccination With Symptomatic SARS-CoV-2 Infection by Time Since Vaccination and Delta Variant Predominance. JAMA 2022; 327:1032–1041.

19. Dickerman BA, Gerlovin H, Madenci AL, et al. Comparative Effectiveness of BNT162b2 and mRNA-1273 Vaccines in U.S. Veterans. N Engl J Med 2022; 386:105–115.

20. De Serres G, Skowronski DM, Wu XW, Ambrose CS. The test-negative design: validity, accuracy and precision of vaccine efficacy estimates compared to the gold standard of randomised placebo-controlled clinical trials. Euro Surveill Bull Eur Sur Mal Transm Eur Commun Dis Bull 2013; 18:20585.

21. Dean NE, Hogan JW, Schnitzer ME. Covid-19 Vaccine Effectiveness and the Test-Negative Design. N Engl J Med 2021; 385:1431–1433.

22. Hitchings MDT, Ranzani OT, Dorion M, et al. Effectiveness of ChAdOx1 vaccine in older adults during SARS-CoV-2 Gamma variant circulation in São Paulo. Nat Commun 2021; 12:6220.

23. Ranzani OT, Hitchings MDT, Dorion M, et al. Effectiveness of the CoronaVac vaccine in older adults during a gamma variant associated epidemic of covid-19 in Brazil: test negative case-control study. The BMJ 2021; 374:n2015.

24. Accorsi EK, Britton A, Fleming-Dutra KE, et al. Association Between 3 Doses of mRNA COVID-19 Vaccine and Symptomatic Infection Caused by the SARS-CoV-2 Omicron and Delta Variants. JAMA 2022; Available at: https://doi.org/10.1001/jama.2022.0470. Accessed 26 January 2022.

25. Schulz WL, Durant TJS, Jr CJT, Hsiao AL, Krumholz HM. Agile Health Care Analytics: Enabling Real-Time Disease Surveillance With a Computational Health Platform. J Med Internet Res 2020; 22:e18707.

26. Khera R, Mortazavi BJ, Sangha V, et al. Accuracy of Computable Phenotyping Approaches for SARS-CoV-2 Infection and COVID-19 Hospitalizations from the Electronic Health Record. MedRxiv Prepr Serv Health Sci 2021; :2021.03.16.21253770.

27. Gangavarapu K, Latif AA, Mullen JL, et al. Outbreak.info genomic reports: scalable and dynamic surveillance of SARS-CoV-2 variants and mutations. 2022; :2022.01.27.22269965. Available at: https://www.medrxiv.org/content/10.1101/2022.01.27.22269965v2. Accessed 4 August 2022.

28. Elbe S, Buckland-Merrett G. Data, disease and diplomacy: GISAID’s innovative contribution to global health. Glob Chall 2017; 1:33–46.

29. Charlson ME, Pompei P, Ales KL, MacKenzie CR. A new method of classifying prognostic comorbidity in longitudinal studies: development and validation. J Chronic Dis 1987; 40:373–383.

30. Kahan BC, Rushton H, Morris TP, Daniel RM. A comparison of methods to adjust for continuous covariates in the analysis of randomised trials. BMC Med Res Methodol 2016; 16:42.

31. Groenwold RHH, Klungel OH, Altman DG, van der Graaf Y, Hoes AW, Moons KGM. Adjustment for continuous confounders: an example of how to prevent residual confounding. CMAJ Can Med Assoc J 2013; 185:401–406.

32. Lipsitch M, Goldstein E, Ray GT, Fireman B. Depletion-of-susceptibles bias in influenza vaccine waning studies: how to ensure robust results. Epidemiol Infect 2019; 147:e306.

33. R Core Team. R: A language and environment for statistical computing. 2021; Available at: https://www.R-project.org/.

34. Seppälä E, Veneti L, Starrfelt J, et al. Vaccine effectiveness against infection with the Delta (B.1.617.2) variant, Norway, April to August 2021. Euro Surveill Bull Eur Sur Mal Transm Eur Commun Dis Bull 2021; 26.

35. Chemaitelly H, Tang P, Hasan MR, et al. Waning of BNT162b2 Vaccine Protection against SARS-CoV-2 Infection in Qatar. N Engl J Med 2021; Available at: https://www.nejm.org/doi/10.1056/NEJMoa2114114. Accessed 7 October 2021.

36. Harder T, Külper-Schiek W, Reda S, et al. Effectiveness of COVID-19 vaccines against SARS-CoV-2 infection with the Delta (B.1.617.2) variant: second interim results of a living systematic review and meta-analysis, 1 January to 25 August 2021. Euro Surveill Bull Eur Sur Mal Transm Eur Commun Dis Bull 2021; 26.

37. Skowronski DM, Febriani Y, Ouakki M, et al. Two-dose SARS-CoV-2 vaccine effectiveness with mixed schedules and extended dosing intervals: test-negative design studies from British Columbia and Quebec, Canada. Clin Infect Dis 2022; :ciac290.

38. Poukka E, Baum U, Palmu AA, et al. Cohort study of Covid-19 vaccine effectiveness among healthcare workers in Finland, December 2020 - October 2021. Vaccine 2022; 40:701–705.

39. Lauring AS, Tenforde MW, Chappell JD, et al. Clinical severity of, and effectiveness of mRNA vaccines against, covid-19 from omicron, delta, and alpha SARS-CoV-2 variants in the United States: prospective observational study. BMJ 2022; 376:e069761.

